# Reliability and validity of clinical tests of cardiorespiratory fitness: A systematic review and meta-analysis

**DOI:** 10.1101/2023.03.08.23286976

**Authors:** Samuel Harkin, Stephen Cousins, Simon Locke, Brett Gordon

## Abstract

**Introduction:** Insufficient physical activity is a significant contributor to non-communicable disease amongst the global population. Insufficient physical activity is directly linked with reduced cardiorespiratory fitness (CRF). CRF is as strong a predictor of mortality as well-established risk-factors such as smoking, hypertension, dyslipidaemia, and type 2 diabetes mellitus, however, it remains the only major risk factor not routinely assessed in primary health care settings. The aim of this review was to assess the validity and reliability of existing submaximal tests of CRF which can be employed in a standard medical consultation for the estimation of CRF and physical function in adults.

**Methods:** A systematic review of the scientific literature was undertaken to find all studies reporting the reliability and/or validity of submaximal tests of CRF and physical function. Studies published up to 12 January 2023 were included in the search of the Medline, Embase, Cinahl, SPORTdiscus, Cochrane library, Informit Health and Web of Science databases. Risk of bias was assessed using the JBI critical appraisal checklist for analytical cross-sectional studies. Data including reliability of the submaximal protocols as measured by test-retest Pearson’s *r* (r) or Intraclass co-efficient (ICC); and validity as measured by the correlation between the submaximal protocol results and the graded exercise test results (r) was extracted. Meta-analyses were performed to determine the overall mean r of the correlation coefficients.

**Results:** In total 1754 studies were identified. Following screening, 143 studies including 15,760 participants were included. All clinical tests included in meta-analysis demonstrated strong reliability. The Siconolfi step test (r=0.81), Incremental shuttle walk test (r=0.768) and 1- minute sit-to-stand test (r=0.65) demonstrated strongest validity following meta-analysis.

**Conclusion:** Based on the validity of the tests outlined, these can be used as an acceptable method of estimating VO2peak in a broad population, without the cost and access issues of formal GXT.

## 1. Introduction

Insufficient physical activity is a significant contributor to non-communicable disease amongst the global population, including obesity, many types of cancer, metabolic syndrome and cardiovascular disease, as well as all-cause mortality (1). In Australia, two thirds of adults report as overweight or obese and over half of Australian adults do not participate in sufficient physical activity to meet the Australian government’s recommended *Physical Activity and Sedentary Behaviour Guidelines for Adults* (2, 3). The economic cost of physical inactivity to the Australian health system is estimated at around $850 million annually, accounting for between 38,400 and 174,000 disability adjusted life years (DALYs) per year (4).

Insufficient physical activity is directly linked with reduced cardiorespiratory fitness (CRF) and reduced physical function (1, 5). Those with low cardiorespiratory fitness face a 70% higher risk of all-cause mortality and a 56% greater risk of cardiovascular disease mortality (5). CRF is as strong a predictor of mortality as well-established risk-factors such as cigarette smoking, hypertension, high cholesterol, and type 2 diabetes mellitus (T2DM) (6). Evidently, measurement of CRF and tracking of progress towards improving CRF should become a standard part of clinical consultations, as is standard of care with hypertension, dyslipidaemia, diabetes monitoring and smoking cessation (6).

Whilst the cost of inactivity is high, there is significant potential for improvement across global populations. CRF can be measured directly via graded exercise testing (GXT) and expressed as maximal oxygen consumption (VO2max), or estimated from peak work rate achieved in GXT (VO2peak) or through submaximal testing via algorithm or correlation. Oxygen consumption can be converted to metabolic equivalents (MET), with most activities having an estimated MET value(7). There is no ‘lower threshold’ for the relative risk reduction benefit of regular exercise (1). For example, a 20% reduction in mortality attributable to cardiac causes is observed for every 1-MET increase in exercise capacity (8). Incidence of falls can be reduced by more than 50% with simple exercise interventions (9). To date, to our knowledge, no study has assessed the validity and reliability of submaximal testing that can be performed in a standard medical consultation, limiting the utility of CRF within clinical consultations.

CRF is recognised as an important marker of functional ability and cardiovascular health; however, it remains the only major risk factor not routinely assessed or regularly monitored in primary health care settings (10). The direct assessment of cardiorespiratory fitness via maximal testing is costly, requires equipment and trained personnel, as well as demanding a maximal effort which is frequently unattained in non-athletic participants (11, 12). Consequently, a large number of submaximal exercise protocols have been developed, involving stationary cycling, running, walking, arm ergometry and stepping; however, time, space or equipment requirements deem many inappropriate for regular medical clinical utility ((12)). Regular and routine clinical testing of physical function and CRF would allow clinicians to determine the CRF and functional capacity of their patients to aid in exercise prescription and counselling, whilst providing the patient with motivation and accountability to improve their health outcomes.

The aim of this review was to assess the validity and reliability of existing submaximal tests of CRF which can be employed in a standard medical consultation for the estimation of CRF and physical function.

## 2. Methods

### 2.1 Review Strategy

A systematic review of the scientific literature was undertaken to find all studies reporting the reliability and/or validity of submaximal tests of CRF and physical function. The study was registered with Prospero registration CRD42022368963 and protocol can be accessed via https://www.crd.york.ac.uk/prospero/display_record.php?RecordID=368963. Studies published up to 12 January 2023 were included in the search of the Medline, Embase, Cinahl, SPORTdiscus, Cochrane library, Informit Health and Web of Science databases . The broad search strategy involved the following terms, limited to English language: ‘Physical function test’ OR ‘exercise test’ OR ‘graded exercise test’ OR ‘GXT’ OR ‘fitness test’ OR ‘squat test’ OR ‘sit to stand’ OR ‘step test’ AND ‘exercise tolerance’ OR ‘exercise capacity’ OR ‘cardiorespiratory fitness’ OR ‘aerobic fitness’ OR ‘aerobic capacity’ OR ‘time to fatigue’ AND ‘validity’ OR ‘valid’ OR ‘reliable’ OR ‘reliability’.

### 2.2 Eligibility criteria

Eligible studies met the following inclusion criteria: (1) English language; (2) published any time from database establishment until 12 January 2023; (3) investigating a clinical test of physical function in (4) participants 18 years of age or older. In order to ensure that studies relevant to the aim of this review were analysed, particularly with regards to tests applicable in a clinical setting, studies were excluded on the following basis: (1) Study type - case reports, not original research, not in English language, conference proceedings; (2) *Clinical* test – equipment requirement beyond scope of that available in a standard Australian medical clinic (e.g. a step, chair or stopwatch), duration greater than ten minutes to administer, expense or technical expertise required to administer; (3) Validity and reliability descriptors – no relevant statistical analysis; (4) Outcomes of interest – no mention of one or more of heart rate, time to fatigue, VO_2_ peak, VO_2_ max, METs, number of repetitions.

### 2.3 Study selection

Studies were selected based on the predetermined inclusion and exclusion criteria. Assessment of study eligibility was performed using Covidence systematic review software (Veritas Health Innovation, Melbourne, Australia, available at www.covidence.org) and conducted independently by two reviewers. Studies were excluded based on title and abstract and the reason for exclusion was recorded, with any disagreement resolved by a third reviewer. Full texts of the remaining studies were assessed by two reviewers, with all authors discussing any disagreements to achieve a consensus view.

### 2.4 Data extraction

One author extracted all relevant data from included studies using a standardised form, with quality control performed on a random sample of 10 papers by two other reviewers. Data extracted included author, year and location of publication; population studied including mean age(standard deviation)[SD], gender split, BMI(SD) and medical condition (where relevant to the study) of participants; inclusion and exclusion criteria; sub-maximal protocol undertaken; graded exercise test undertaken; reliability and validity statistics; and other outcome measures of note (HR, VO_2_peak, RPE, 6MWD, sit-to-stand repetitions). The primary outcomes of interest were reliability of the submaximal protocols as measured by test-retest Pearson’s *r* (r) or Intraclass co-efficient (ICC); and validity as measured by the correlation between the submaximal protocol results and the graded exercise test results (r).

### 2.5 Risk of bias assessment

The risk of bias of included studies was assessed using the JBI critical appraisal checklist for analytical cross sectional studies (JBIC) (13). This checklist assesses specific domains of the studies to determine the potential risk of bias that can be answered with yes, no, or unclear. If the answer was yes, the question was assigned a score of 1. If the answer was no, unclear, or not applicable, it was assigned a score of 0. Studies with a score of 7-8 were deemed low risk of bias, 4-6 moderate and 0-3 high. No studies were excluded on the basis of risk of bias assessment.

### 2.6 Data synthesis

Meta-analyses were performed to determine the overall mean r of the correlation coefficients. Data including reliability of the submaximal protocols as measured by test-retest Pearson’s *r* (r) or Intraclass co-efficient (ICC); and validity as measured by the correlation between the submaximal protocol results and the graded exercise test results (r), was pooled using a random-effects model. Jamovi Version 2.2 (Computer Software retrieved from https://www.jamovi.org) was used for statistical analysis.

## 3. Results

### 3.1 Paper identification

In total 1754 studies (title and abstract) were identified following deletion of duplicates (n=80). Following screening, 1466 were excluded, with 283 full texts obtained for further eligibility assessment. 143 full-text papers were included for review *(Figure 1)*.

**Fig 1:**
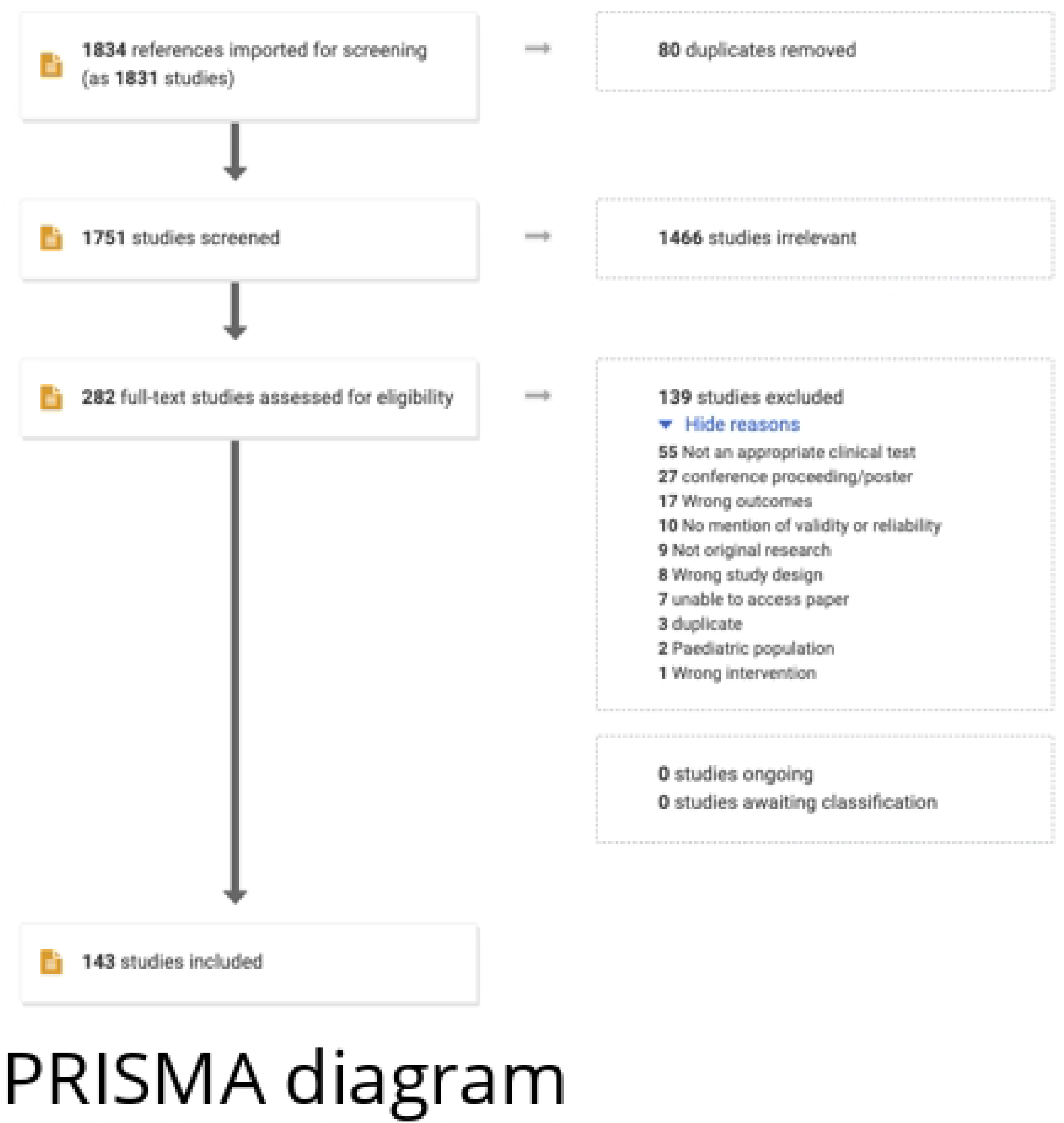
PRISMA diagram of included studies

### 3.2 Sub-maximal test characteristics

The 143 included papers studied 49 different clinical tests of physical function. 75 studies assessed the 6-minute walk test (6MWT) (14, 15, 16, 17, 18, 19, 20, 21, 22, 23, 24, 25, 26, 27, 28, 29, 30, 31, 32, 33, 34, 35, 36, 37, 38, 39, 40, 41, 42, 43, 44, 45, 46, 47, 48, 49, 50, 51, 52, 53, 54, 55, 56, 57, 58, 59, 60, 61, 62, 63, 64, 65, 66, 67, 68, 69, 70, 71, 72, 73, 74, 75, 76, 77, 78, 79, 80, 81, 82, 83, 84, 85, 86, 87). Fifteen studies analysed the incremental shuttle walk test (ISWT) (18, 37, 56, 58, 68, 88, 89, 90, 91, 92, 93, 94, 95, 96, 97). The 1-minute sit to stand(1mSTS) was studied in eight papers, the 6-minute step test (6MST) in seven, ‘timed up and go test’ (TUGT) in six, and the Siconolfi step test (SST) in five (70, 80, 86, 87, 98, 99, 100, 101, 102, 103, 104, 105, 106, 107, 108, 109, 110, 111, 112, 113, 114). Seven tests were studied in three papers, five tests were studied twice and fifty once. A degree of heterogeneity existed amongst the exact test protocols for many of the clinical tests described by the same name, however they were extracted as named by the authors.

### 3.3 Graded exercise test characteristics

A graded exercise test was included in 71 of the 143 studies included. Of these, 38 included a cycle ergometry based graded exercise test, with one additional study recumbent cycle ergometry. There were 32 treadmill based graded exercise tests included.

### 3.4 Quality assessment

Table 1 provides a summary of how each individual paper rated per the JBI critical appraisal checklist. Fifty (34.9%) included studies scored 7 or 8 to be deemed low risk of bias. Eighty-three (58.2%) had a moderate risk, and 10 (6.9%) high risk of bias.

**Table 1:** Quality assessment JBI critical appraisal checklist.

| Study ID | Q1 | Q2 | Q3 | Q4 | Q5 | Q6 | Q7 | Q8 | Total |
| --- | --- | --- | --- | --- | --- | --- | --- | --- | --- |
| Cheng 2020 | Yes | Yes | Yes | Yes | Yes | Yes | Yes | Yes | 8 |
| Barbosa 2022 | Yes | Yes | Yes | Yes | Yes | Yes | Yes | Yes | 8 |
| Cooney 2011 | Yes | Yes | Yes | Yes | Yes | Yes | Yes | Yes | 8 |
| Cooney 2013 | Yes | Yes | Yes | Yes | Yes | Yes | Yes | Yes | 8 |
| Crook 2017 | Yes | Yes | Yes | Yes | Yes | Yes | Yes | Yes | 8 |
| DeCamargo 2013 | Yes | Yes | Yes | Yes | Yes | Yes | Yes | Yes | 8 |
| Bonnevie 2019 | Yes | Yes | Yes | Yes | Yes | Yes | Yes | Yes | 8 |
| Brinklov 2016 | Yes | Yes | Yes | Yes | Yes | Yes | Yes | Yes | 8 |
| Granger 2015 | Yes | Yes | Yes | Yes | Yes | Yes | Yes | Yes | 8 |
| Reed 2020 | Yes | Yes | Yes | Yes | Yes | Yes | Yes | Yes | 8 |
| Grosbois 2016 | Yes | Yes | Yes | Yes | Yes | Yes | Yes | Yes | 8 |
| Guazzi 2009 | Yes | Yes | Yes | Yes | Yes | Yes | Yes | Yes | 8 |
| Hansen 2018 | Yes | Yes | Yes | Yes | Yes | Yes | Yes | Yes | 8 |
| Maldonado-Martin 2006 | Yes | Yes | Yes | Yes | Yes | Yes | Yes | Yes | 8 |
| Manali 2010 | Yes | Yes | Yes | Yes | Yes | Yes | Yes | Yes | 8 |
| Shulman 2019 | Yes | Yes | Yes | Yes | Yes | Yes | Yes | Yes | 8 |
| Vancampfort 2016 | Yes | Yes | Yes | Yes | Yes | Yes | Yes | Yes | 8 |
| Lee 2015 | Yes | Yes | Yes | Yes | Yes | Yes | Yes | Yes | 8 |
| Freene 2021 | Yes | Yes | Yes | N/A | Yes | Yes | Yes | Yes | 7 |
| Tremblay-Labrecque 2020 | Yes | Yes | Yes | Yes | Yes | No | Yes | Yes | 7 |
| Munari 2021 | Yes | Yes | Yes | Yes | Yes | No | Yes | Yes | 7 |
| Marinho 2021 | Yes | Yes | Yes | Yes | Yes | No | Yes | Yes | 7 |
| Tsuji 2022 | Yes | Yes | Yes | Yes | Yes | No | Yes | Yes | 7 |
| DalCorso 2012 | Yes | Yes | Yes | Yes | Yes | No | Yes | Yes | 7 |
| DiThommazo-Luporini 2015 | Yes | Yes | Yes | N/A | Yes | Yes | Yes | Yes | 7 |
| Lee 2018 | Yes | Yes | Yes | Yes | Yes | No | Yes | Yes | 7 |
| Beckerman 2019 | Yes | Yes | Yes | Unclear | Yes | Yes | Yes | Yes | 7 |
| Beekman 2013 | Yes | Yes | No | Yes | Yes | Yes | Yes | Yes | 7 |
| Buch 2007 | Yes | Yes | Yes | Yes | Yes | No | Yes | Yes | 7 |
| Fowler 2011 | Yes | Yes | Yes | Yes | Yes | No | Yes | Yes | 7 |
| Gayda 2003 | Yes | Yes | Yes | Yes | Yes | No | Yes | Yes | 7 |
| Gomberg-Maitland 2007 | Yes | Yes | Yes | Yes | Yes | No | Yes | Yes | 7 |
| OzcanKahraman 2020 | Yes | Yes | Yes | Yes | Yes | No | Yes | Yes | 7 |
| Peloquin 1998 | Yes | Yes | Yes | Yes | Yes | No | Yes | Yes | 7 |
| PereiradeSousa 2008 | Yes | Yes | Yes | Yes | Yes | No | Yes | Yes | 7 |
| Radtke 2017 | Yes | Yes | Yes | Yes | Yes | No | Yes | Yes | 7 |
| Hansen 2016 | Yes | Yes | Yes | No | Yes | Yes | Yes | Yes | 7 |
| Hornby 2019 | Yes | Yes | Yes | Yes | Yes | No | Yes | Yes | 7 |
| Kehmeier 2016 | Yes | Yes | Yes | Yes | Yes | No | Yes | Yes | 7 |
| Keren 1980 | Yes | Yes | Yes | N/A | Yes | Yes | Yes | Yes | 7 |
| Leung 2006 | Yes | Yes | Yes | No | Yes | Yes | Yes | Yes | 7 |
| Metz 2018 | Yes | Yes | Yes | Yes | Yes | No | Yes | Yes | 7 |
| Muller 2015 | Yes | No | Yes | Yes | Yes | Yes | Yes | Yes | 7 |
| Nielsen 1997 | Yes | Yes | Yes | Yes | No | Yes | Yes | Yes | 7 |
| Sartor 2016 | Yes | Yes | Yes | N/A | Yes | Yes | Yes | Yes | 7 |
| Schmidt 2013 | Yes | Yes | Yes | Yes | Yes | No | Yes | Yes | 7 |
| Scivoletto 2011 | Yes | Yes | Yes | Yes | Yes | No | Yes | Yes | 7 |
| Simonsick 2006 | Yes | Yes | Yes | No | Yes | Yes | Yes | Yes | 7 |
| Tarrant 2020 | Yes | Yes | Yes | Yes | Yes | No | Yes | Yes | 7 |
| Vancampfort 2015 | Yes | No | Yes | Yes | Yes | Yes | Yes | Yes | 7 |
| Kakitsuka 2020 | Yes | Yes | Yes | N/A | Yes | No | Yes | Yes | 6 |
| Ang 2022 | Yes | Yes | Yes | N/A | Yes | No | Yes | Yes | 6 |
| Vancampfort 2021 | Yes | No | No | Yes | Yes | Yes | Yes | Yes | 6 |
| Diaz-Balboa 2022 | No | Yes | Yes | Yes | Yes | No | Yes | Yes | 6 |
| Gephine 2020 | Yes | Yes | Yes | Yes | No | No | Yes | Yes | 6 |
| daCunha-Filho 2007 | Yes | Yes | Yes | Yes | No | No | Yes | Yes | 6 |
| DalCorso 2007 | Yes | Yes | Yes | No | Yes | No | Yes | Yes | 6 |
| Kervio 2003 | Yes | Yes | Yes | N/A | Yes | No | Yes | Yes | 6 |
| Kervio 2004 | Yes | Yes | Yes | Yes | No | No | Yes | Yes | 6 |
| Knight 2014 | Yes | Yes | Yes | N/A | Yes | No | Yes | Yes | 6 |
| Lemanska 2019 | Yes | Yes | Yes | Yes | No | No | Yes | Yes | 6 |
| Asakuma 1999 | Yes | Yes | Yes | Yes | No | No | Yes | Yes | 6 |
| Carbonell-Baeza 2015 | Yes | Yes | Yes | Yes | No | No | Yes | Yes | 6 |
| Carvalho 2011 | Yes | Yes | Yes | Yes | No | No | Yes | Yes | 6 |
| Giacomantonio 2020 | Yes | Yes | Yes | Yes | No | No | Yes | Yes | 6 |
| Olper 2011 | Yes | Yes | No | Yes | Yes | Yes | No | Yes | 6 |
| Pankoff 2000 | Yes | Yes | Yes | No | Yes | No | Yes | Yes | 6 |
| PV@loquin 1998 | Yes | Yes | Yes | Yes | No | No | Yes | Yes | 6 |
| Radtke 2016 | Yes | No | Yes | Yes | Yes | No | Yes | Yes | 6 |
| Reilly 2010 | Yes | No | Yes | N/A | Yes | Yes | Yes | Yes | 6 |
| Guo 2018 | Yes | Yes | Yes | N/A | Yes | No | Yes | Yes | 6 |
| Hwang 2016 | Yes | No | Yes | Yes | Yes | No | Yes | Yes | 6 |
| Irisawa 2014 | Yes | Yes | Yes | Yes | No | No | Yes | Yes | 6 |
| Jehn 2009 | Yes | Yes | No | Yes | Yes | No | Yes | Yes | 6 |
| Jette 1992 | Yes | Yes | Unclear | Yes | Yes | No | Yes | Yes | 6 |
| Lin 2008 | Yes | Yes | No | Yes | Yes | No | Yes | Yes | 6 |
| Magalhaes 2020 | Yes | Yes | Yes | Yes | No | No | Yes | Yes | 6 |
| Mazzoni 2018 | Yes | Yes | Yes | Yes | No | No | Yes | Yes | 6 |
| Mercer 1998 | Yes | Yes | Yes | Yes | No | No | Yes | Yes | 6 |
| Modai 2015 | Yes | Yes | Yes | No | Yes | No | Yes | Yes | 6 |
| Reychler 2018 | No | Yes | Yes | Yes | Yes | No | Yes | Yes | 6 |
| Rodrigues 2016 | Yes | Yes | Unclear | Yes | Yes | No | Yes | Yes | 6 |
| Simmonds 2002 | Yes | Yes | Yes | Yes | No | No | Yes | Yes | 6 |
| Spagnuolo 2010 | Yes | Yes | No | Yes | Yes | No | Yes | Yes | 6 |
| Sykes 2004 | Yes | No | Yes | N/A | Yes | Yes | Yes | Yes | 6 |
| Vancampfort 2020 | Yes | No | Yes | No | Yes | Yes | Yes | Yes | 6 |
| Vancampfort 2011 | Yes | Yes | Yes | No | Yes | No | Yes | Yes | 6 |
| Webb 2014 | No | Yes | Yes | Yes | Yes | No | Yes | Yes | 6 |
| Zanini 2015 | Yes | Yes | No | Yes | Yes | No | Yes | Yes | 6 |
| Lazaro-Martinez 2022 | No | Yes | Yes | Yes | No | No | Yes | Yes | 5 |
| Quintino 2021 | Yes | No | Yes | No | Yes | No | Yes | Yes | 5 |
| Vilarinho 2022 | Yes | Yes | No | Yes | Yes | No | No | Yes | 5 |
| Chow 2022 | Yes | Yes | No | Yes | No | No | Yes | Yes | 5 |
| Cox 1992 | No | Yes | Yes | No | Yes | No | Yes | Yes | 5 |
| Curb 2006 | Yes | No | No | No | Yes | Yes | Yes | Yes | 5 |
| Eiser 2003 | Yes | Yes | No | No | Yes | No | Yes | Yes | 5 |
| Kierkegaard 2007 | Yes | Yes | No | Yes | No | No | Yes | Yes | 5 |
| Kieu 2020 | Yes | Yes | Yes | No | No | No | Yes | Yes | 5 |
| Kon 2013 | Yes | No | No | Yes | Yes | No | Yes | Yes | 5 |
| Bardin 2012 | Yes | Yes | No | No | Yes | Unclear | Yes | Yes | 5 |
| BenSaad 2009 | Yes | Yes | No | Yes | No | No | Yes | Yes | 5 |
| Beretta 2007 | Yes | Yes | No | Yes | Yes | No | No | Yes | 5 |
| Boer 2016 | Yes | Yes | Unclear | No | Yes | No | Yes | Yes | 5 |
| Bronner 2014 | Yes | Yes | N/A | No | Yes | No | Yes | Yes | 5 |
| Hong 2019 | No | Yes | Yes | N/A | Yes | No | Yes | Yes | 5 |
| Jones 2018 | Yes | Yes | No | Yes | No | No | Yes | Yes | 5 |
| Keisuke 2015 | Yes | Yes | Yes | N/A | No | No | Yes | Yes | 5 |
| Mänttari 2018 | Yes | No | Yes | N/A | No | Yes | Yes | Yes | 5 |
| Marcora 2007 | Yes | Yes | Yes | Unclear | No | No | Yes | Yes | 5 |
| Mossberg 2003 | Yes | Yes | No | No | Yes | No | Yes | Yes | 5 |
| Nathan 2015 | No | Yes | No | Yes | Yes | Yes | No | Yes | 5 |
| Rolland 2004 | Yes | No | Yes | N/A | Yes | Yes | No | Yes | 5 |
| Santo 2003 | Yes | No | Yes | N/A | Yes | No | Yes | Yes | 5 |
| Vagaggini 2003 | Yes | Yes | Yes | No | No | No | Yes | Yes | 5 |
| deJesus 2022 | Unclear | Yes | Yes | No | No | No | Yes | Yes | 4 |
| Correa 2013 | Yes | No | No | Yes | No | No | Yes | Yes | 4 |
| Kohlbrenner 2020 | Yes | Yes | No | Unclear | No | No | Yes | Yes | 4 |
| Bauman 1997 | Yes | Yes | No | No | No | Unclear | Yes | Yes | 4 |
| Berghmans 2013 | Yes | No | Yes | No | Yes | No | Unclear | Yes | 4 |
| Beutner 2015 | No | No | Yes | No | Yes | No | Yes | Yes | 4 |
| Borel 2010 | No | No | Yes | No | Yes | No | Yes | Yes | 4 |
| Cabeza-Ruiz 2019 | Yes | No | Yes | No | No | No | Yes | Yes | 4 |
| Carter 2002 | Yes | No | Yes | Yes | No | No | No | Yes | 4 |
| Francis 1988 | No | Yes | Yes | N/A | No | No | Yes | Yes | 4 |
| Hanson 2012 | No | No | Yes | Unclear | Yes | Yes | No | Yes | 4 |
| Levesque 2019 | Yes | Yes | No | Yes | No | No | No | Yes | 4 |
| Ljungquist 2003 | Yes | No | No | No | Yes | No | Yes | Yes | 4 |
| Siconolfi 1985 | Yes | No | Yes | N/A | No | No | Yes | Yes | 4 |
| Thornton 2017 | No | No | Yes | N/A | Yes | No | Yes | Yes | 4 |
| Wanwisa 2015 | No | No | Yes | Unclear | Yes | No | Yes | Yes | 4 |
| Witham 2012 | No | No | Yes | No | Yes | No | Yes | Yes | 4 |
| deMelo 2022 | No | Yes | No | N/A | No | No | Yes | Yes | 3 |
| Aadahl 2013 | Yes | No | No | No | Unclear | No | Yes | Yes | 3 |
| Chatterjee 2005 | No | No | Yes | N/A | No | No | Yes | Yes | 3 |
| Kristjánsdóttir 2004 | No | No | Yes | No | No | No | Yes | Yes | 3 |
| Grant 1999 | No | No | Yes | N/A | No | No | Yes | Yes | 3 |
| Montgomery 1992 | No | No | No | No | Yes | No | Yes | Yes | 3 |
| Selig 2000 | No | No | Yes | No | No | No | Yes | Yes | 3 |
| Donaldson 2019 | No | No | No | No | No | No | Yes | Yes | 2 |
| Benton 2009 | No | No | No | No | Unclear | Unclear | Yes | Yes | 2 |
| VanGraan 1970 | No | No | Yes | N/A | No | No | Unclear | No | 1 |

### 3.4 Participant characteristics

In total 15,670 participants were included across the 143 studies. The smallest study included 5 participants (39), the largest 5287 (115). Six studies reported no sex distribution, of the studies reporting, 5588 participants were female (approximately 37%).

#### 3.4.1 Special populations

Thirty-nine studies included no specific medical condition inclusion criteria. Participants selected for respiratory conditions (including COPD, restrictive lung disease and sleep apnoea) were studied in 29 papers. Cardiovascular conditions were the focus in 21. Musculoskeletal, neurological, auto-immune, psychological and age based sub-groups were also studied in the remaining papers.

### 3.5 Measures

All included studies reported clinical test performance correlation with GXT performance in terms of Pearson’s r. Test-retest reliability was reported in terms of ICC or Pearson’s r.

### 3.6 Clinical tests

#### 3.6.1 Walk test protocols

##### 3.6.1.1 6MWT

A total of 71 included papers studied the 6MWT, with 21 cohorts from a total of 19 studies meeting inclusion in the meta-analysis for validity with correlation between 6MWT and directly measure VO2 peak on GXT results included. In total these studies included 1836 participants with a broad range of medical conditions. Overall the 6MWT demonstrated moderate positive correlation with performance on the graded exercise test (GXT), with r=0.581 (figure 2). The largest study, Shulman et al studied 574 adults older than 40 years awaiting elective non-cardiac surgery, and all participants had at least one cardiac risk factor, with low positive correlation overall r=0.36 (28). Kervio et al’s study of 24 patients of mean age 65 with NYHA grade II or III congestive cardiac failure (CCF) found the strongest association with GXT performance r=0.8 (16). Granger et al’s study of a cohort of 20 male lung cancer patients found the lowest correlation, r = 0.24 (58). Pankoff et al’s study of a fibromyalgia cohort pre and post exercise program found the pre-exercise cohort correlation of r=0.33 (30). The only included study of a ‘healthy’ population, Hong et al, involved a cohort of 73 healthy adults (37male, 36 female), mean age 30.8 who were screened for cardiorespiratory, orthopaedic and musculoskeletal conditions prior to participation, and found a correlation between 6MWT and GXT performance of r=0.671. Overall four studies of five CCF populations were included, with correlations ranging from 0.54 to 0.8 (16, 17, 23, 25). Two studies analysed pulmonary artery hypertension populations, with strong correlation noted r=0.77 and r=0.72 (18, 19).

**Fig 2:**
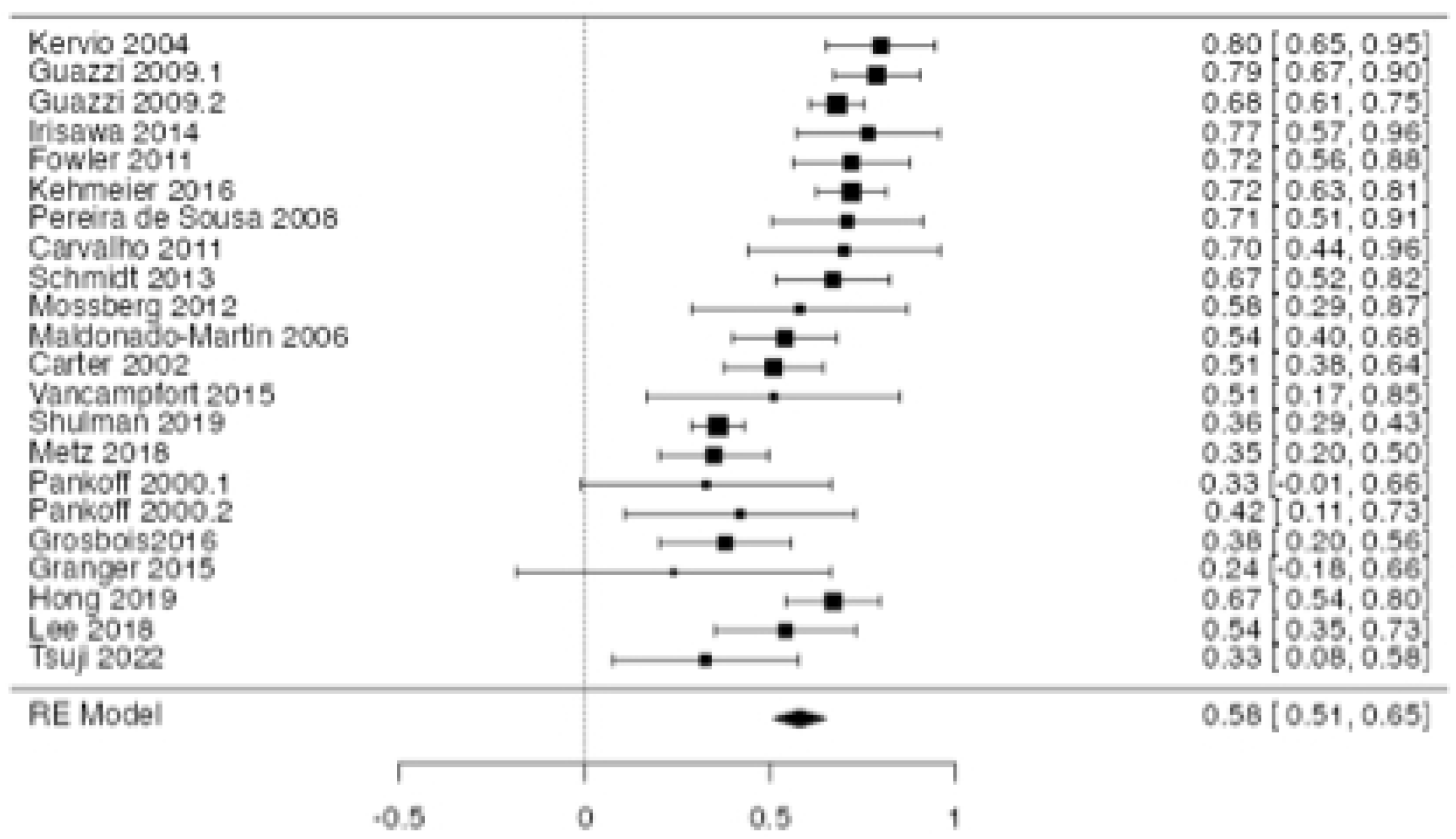
6MWT performance vs GXT correlation forest plot

**Fig 3:**
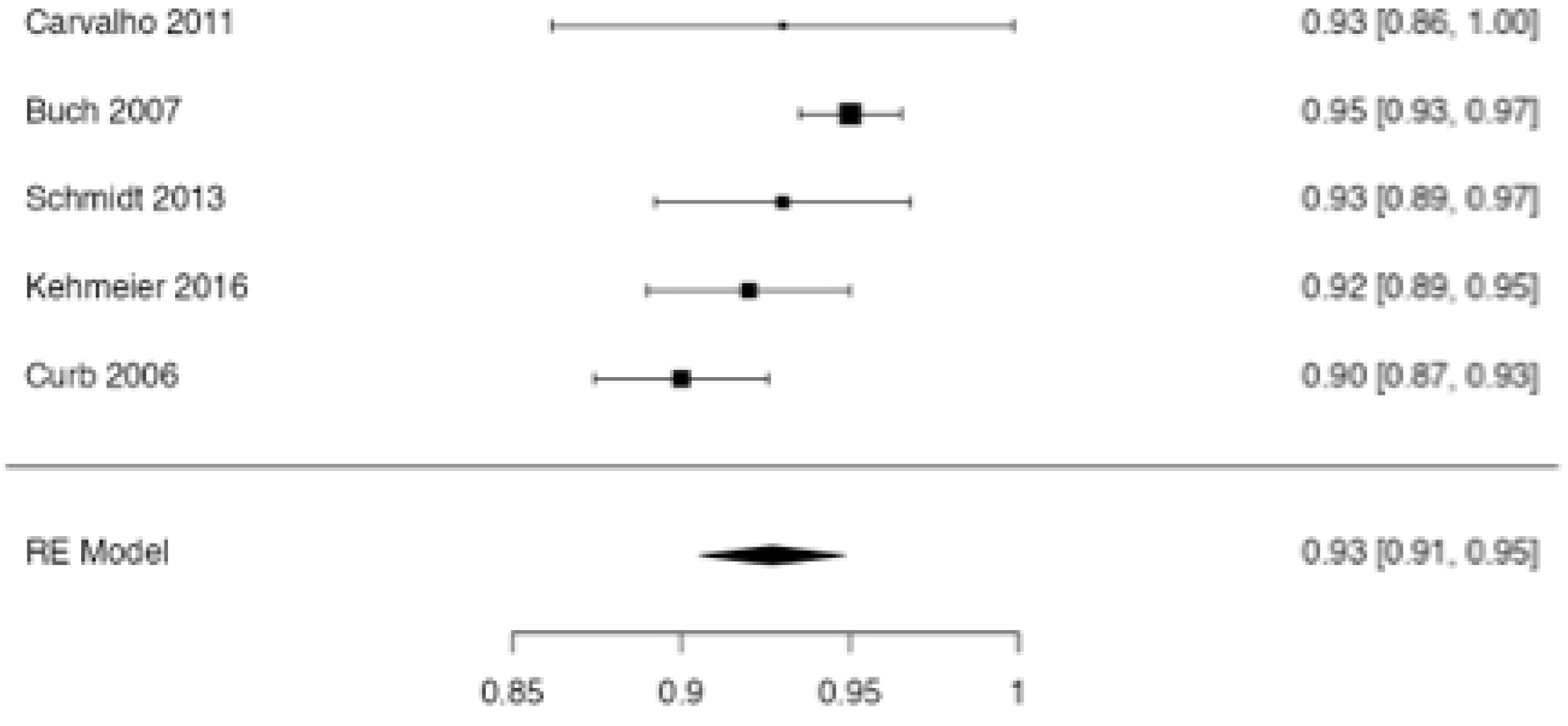
6MWT reliability (pearson’s r) forest plot

**Fig 4:**
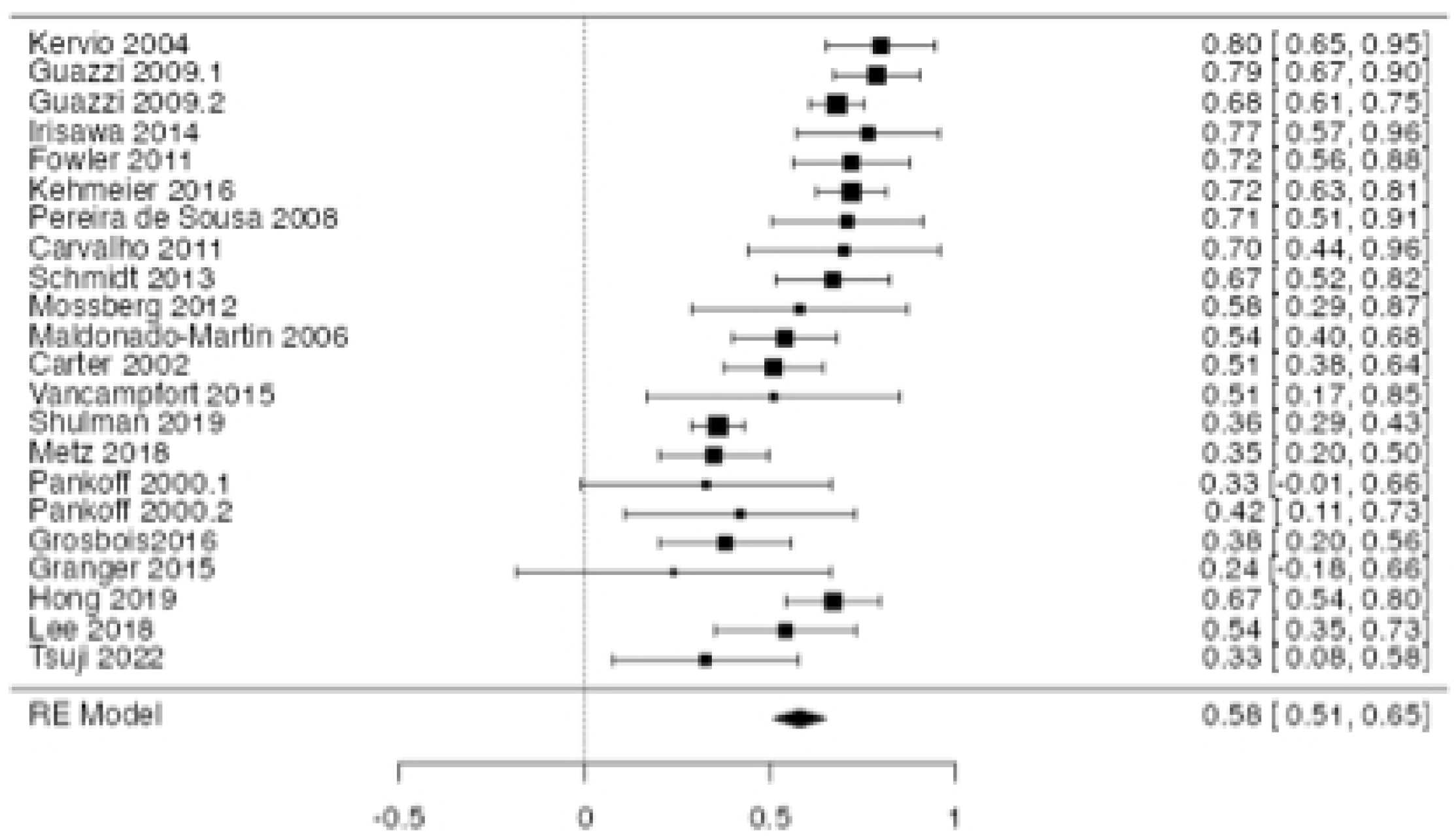
6MWT reliability (ICC) forest plot

**Fig 5:**
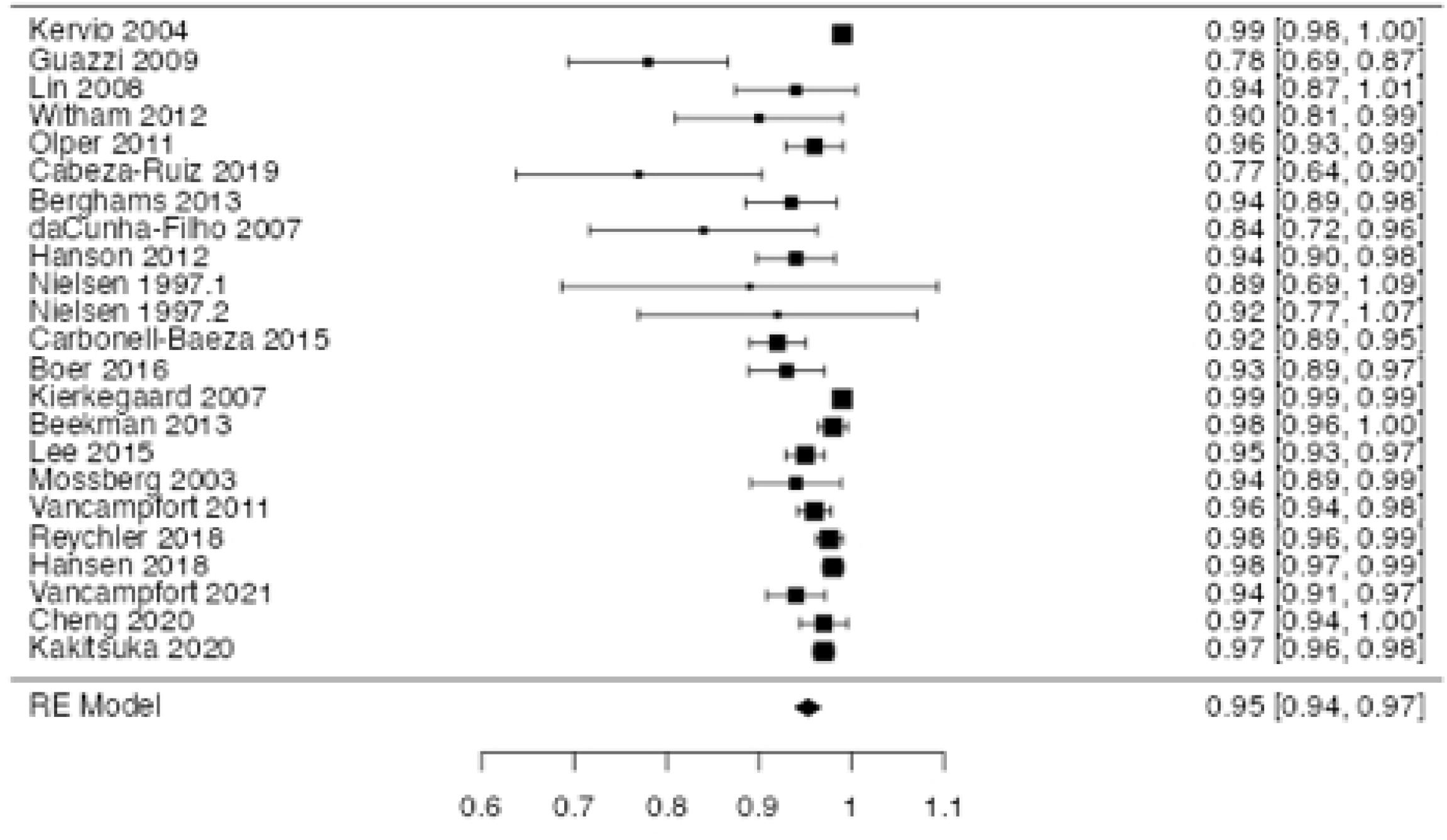
ISWT performance vs GXT correlation forest plot

**Fig 6:**
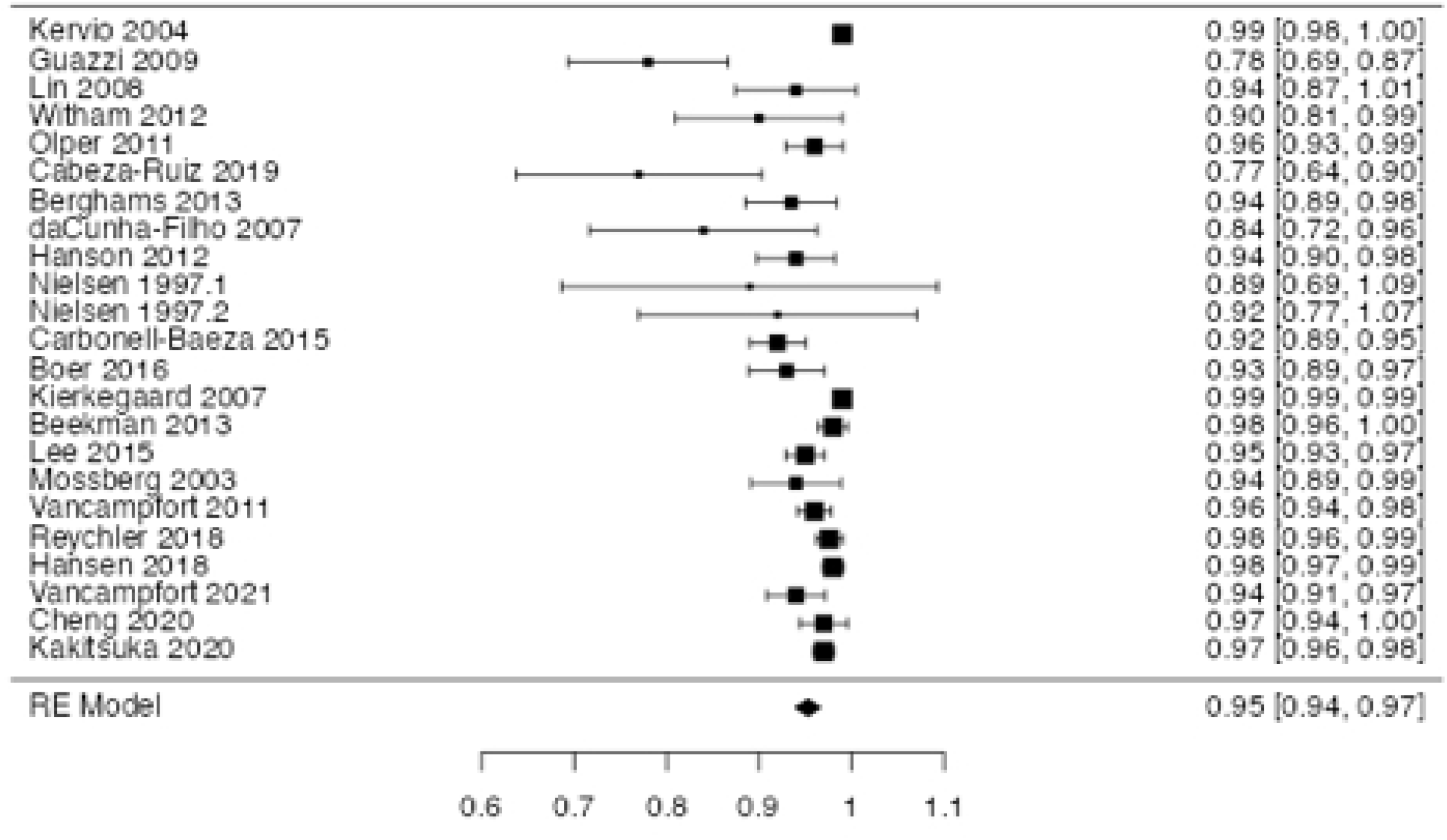
ISWT reliability (ICC) forest plot

**Fig 7:**
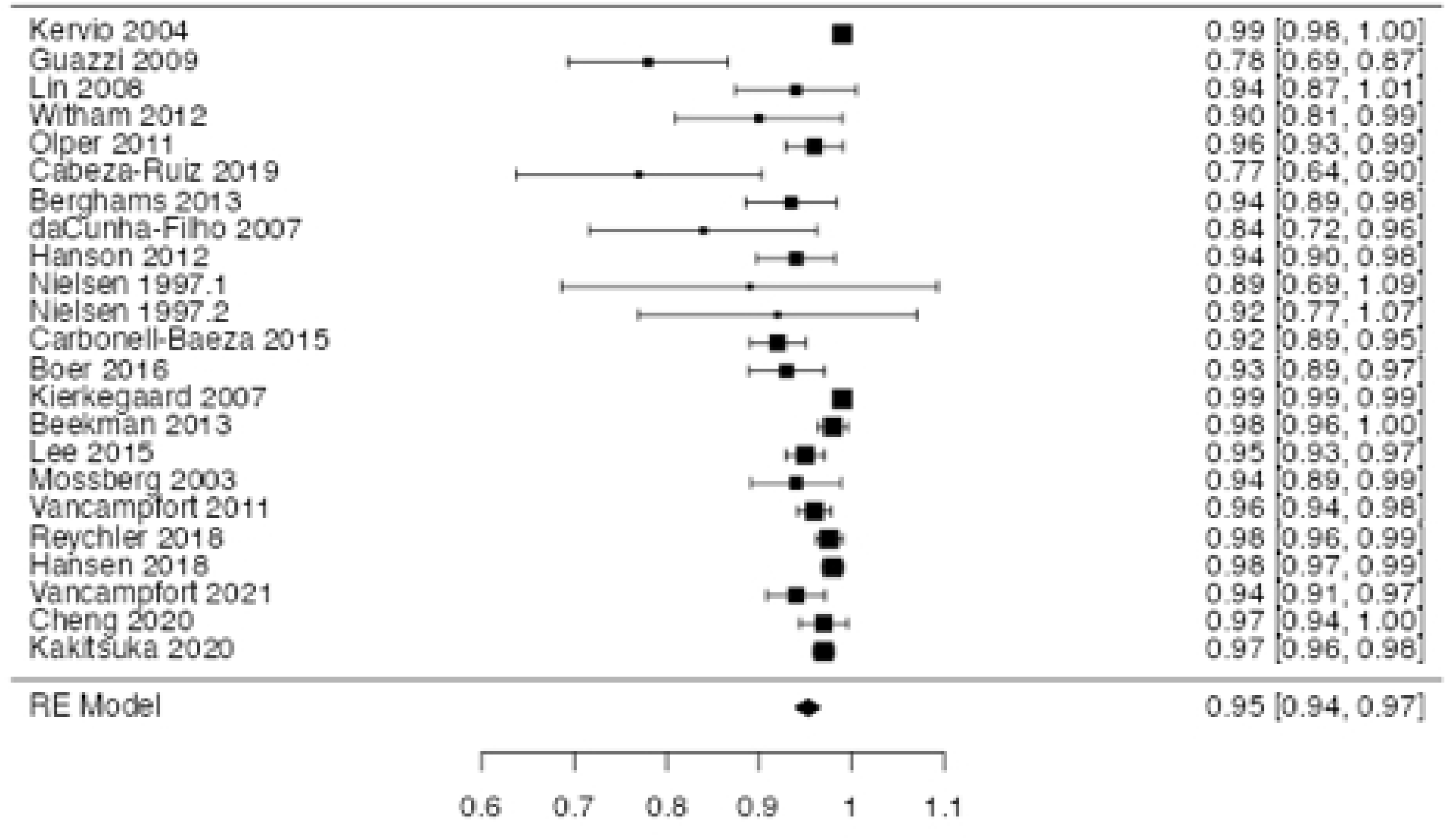
SST vs GXT correlation forest plot

**Fig 8:**
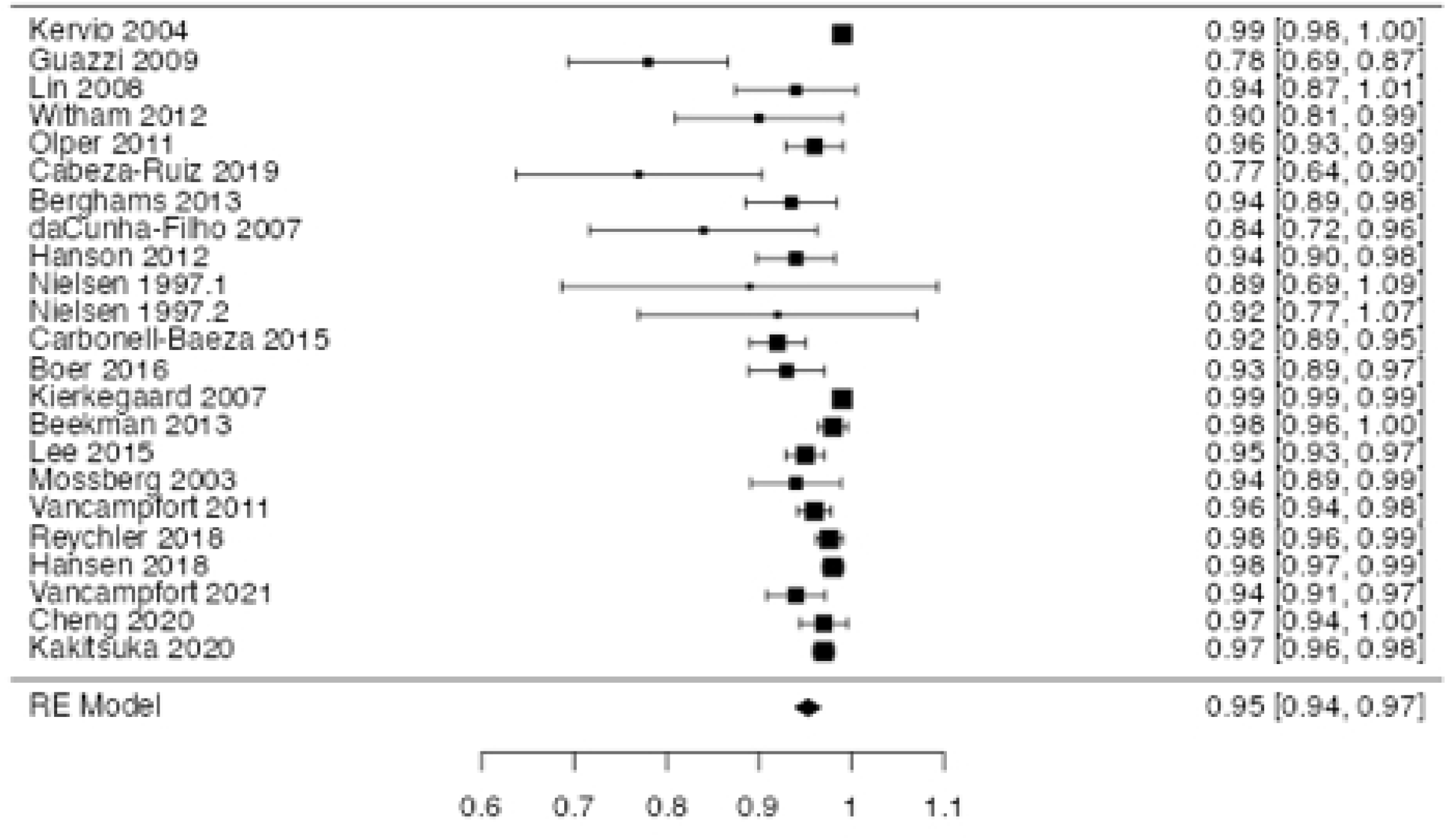
SST reliability (ICC) forest plot

Test-retest reliability of the 6MWT was studied in 27 studies of 1506 participants. In meta-analysis of 22 studies of 965 patients, with reliability measured by ICC, an overall ICC of 0.95, indicating strong test-retest reliability. For the remaining five studies of 541 participants r=0.93.

##### 3.6.1.2 ISWT

Fourteen studies included the ISWT, with 5 studies meeting inclusion in the meta-analysis for validity (18, 58, 88, 92, 93, 94). In total these studies included 114 participants. Overall the ISWT demonstrated strong positive correlation with performance on GXT, with r=0.768. De Camargo et al’s study of a cohort of 75 patients with bronchiectasis was the largest cohort, with r=0.72 compared with a cycle ergometer GXT (88). Barbosa et al’s study of 50 asthma patients demonstrated the strong validity of the ISWT in this cohort at r=0.9 (93). Irisawa et al’s study of 19 patients with pulmonary artery hypertension (PAH) also demonstrated the strong validity, r=0.866, with participants recording the lowest mean ISWT distance (ISWD) at 359.4m, compared with 441m and 410m for the de Camargo and Granger cohorts respectively (18, 58, 88). Granger et al studied an all-male population of 20 non-small cell lung cancer patients with r=0.61 (58).

Test-retest reliability was very high across the five studies meeting inclusion criteria for reliability, with ICC=0.94 from 284 participants (37, 63, 88, 93, 94).

##### 3.6.1.3 2-minute walk test (2MWT)

In total four papers included the 2MWT, however an insufficient number met the inclusion criteria to be meta-analysed. Two studies assessed the 2MWT performance correlation with GXT. Beckerman et al studied 141 patients with multiple sclerosis, finding poor validity r=0.44 (116). Leung et al studied 45 patients with moderate to severe COPD, finding a moderate correlation r=0.56 (66).

Two studies found strong test-retest reliability for the 2MWT, with again insufficient numbers included to be meta-analysed. Vancampfort (66) and Leung respectively found ICC=0.96 and ICC=0.99 (66, 83).

#### 3.6.2 Step tests

##### 3.6.2.1 Siconolfi step test (SST)

Three studies met the inclusion criteria for validity meta-analysis, with an overall population of 138 patients included and a strong correlation of r=0.81 established. Lemanska et al’s study of 66 men with prostate cancer was the largest included, with the weakest correlation at r=0.69 (112). Siconolfi’s original study of 48 healthy adults found a correlation of r=0.92 with a cycle ergometer GXT (114). Cooney et al found a strong correlation of r = 0.79 in 24 patients with rheumatoid arthritis (RA)(111).

Reliability was very strong for the SST across 120 patients from 3 studies, with ICC = 0.92 demonstrating good test-retest reliability (111, 112, 113).

##### 3.6.2.2 Chester step test (CST)

Two included studies assessed the validity of the Chester step test (117, 118). Sykes et al assessed 68 healthy adults and found strong correlation between CST and GXT result, r=0.92, with a standard error of predicted estimate of aerobic capacity (VO2peak) 3.9mlO2/kg/min (118). Reed et al in their 2020 study of 47 cardiac rehabilitation participants also found high moderate correlation r=0.693 (117). Their CST involved adjustment of step height within a range between 15-30cm “suitable to participants functional level”. Of note the Sykes population achieved a mean VO2Max of 52.1mlo2/kg/min, suggesting a good level of CRF, and validity in a population with good levels of fitness.

Neither study included correlation reliability statistics. Sykes et al (119) reported good test-retest reliability using the Bland and Altman method, finding a mean difference of - 0.7mlO2/kg/min (119).

##### 3.6.2.3 6 minute step test (6MST)

Two studies correlated GXT performance with 6MST performance to determine validity, Giacomantonio et al found a strong correlation with GXT of r=0.88 in a population of 28 participants with two or more CVD risk factors, whereas Marinho et al found a weaker r=0.59 in 27 heart failure with reduced ejection fraction (HFrEF) patients (64, 109). Five studies of a total of 194 participants found good reliability, ICC = 0.97 across varied populations of healthy adults, an obstructive sleep apnoea cohort, patients with COPD and the above CVD and HFrEF cohort (Figure 9) (64, 72, 105, 108, 109).

**Figure 9:**
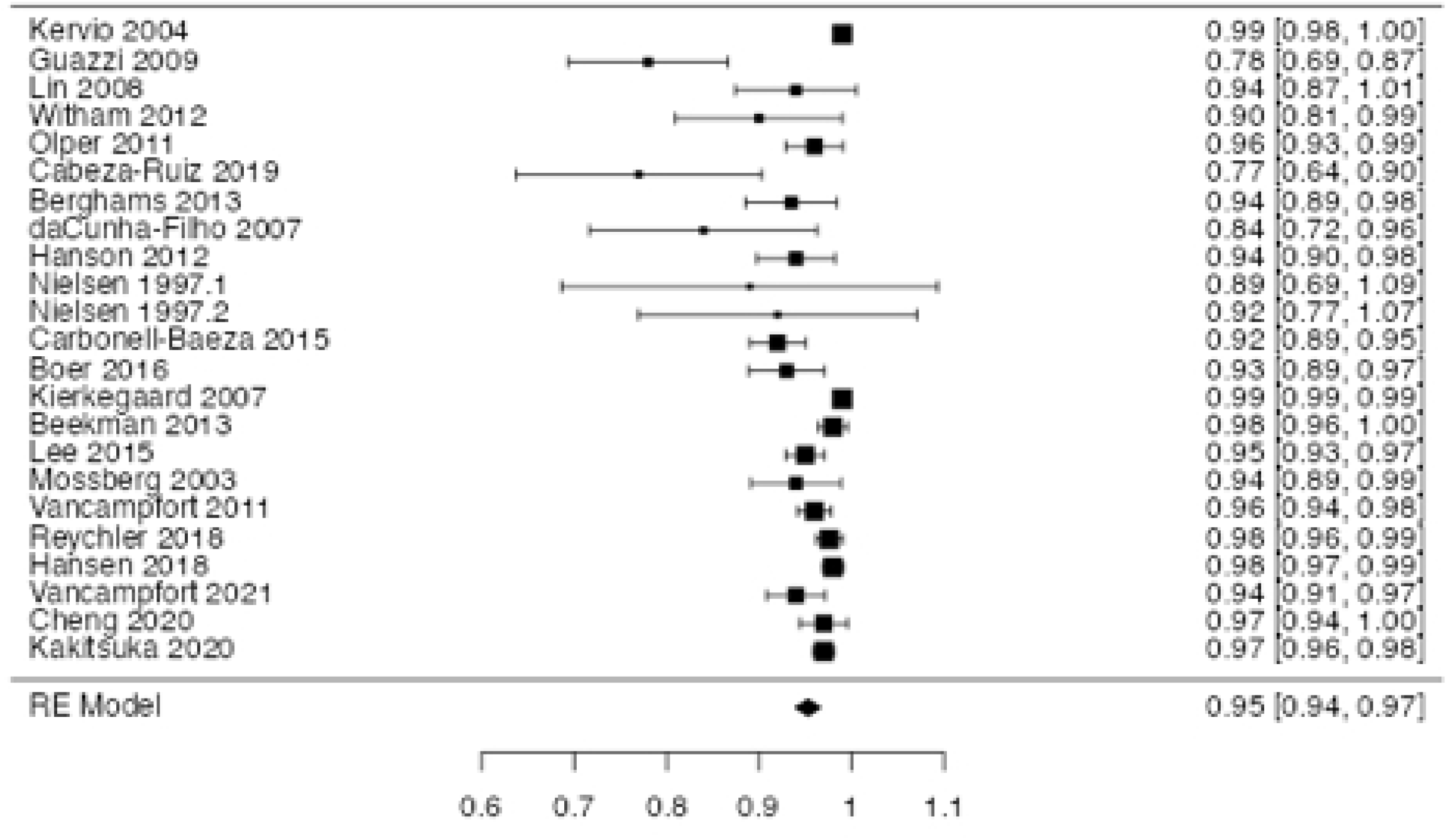
6MST reliability (ICC) forest plot

**Fig 10:**
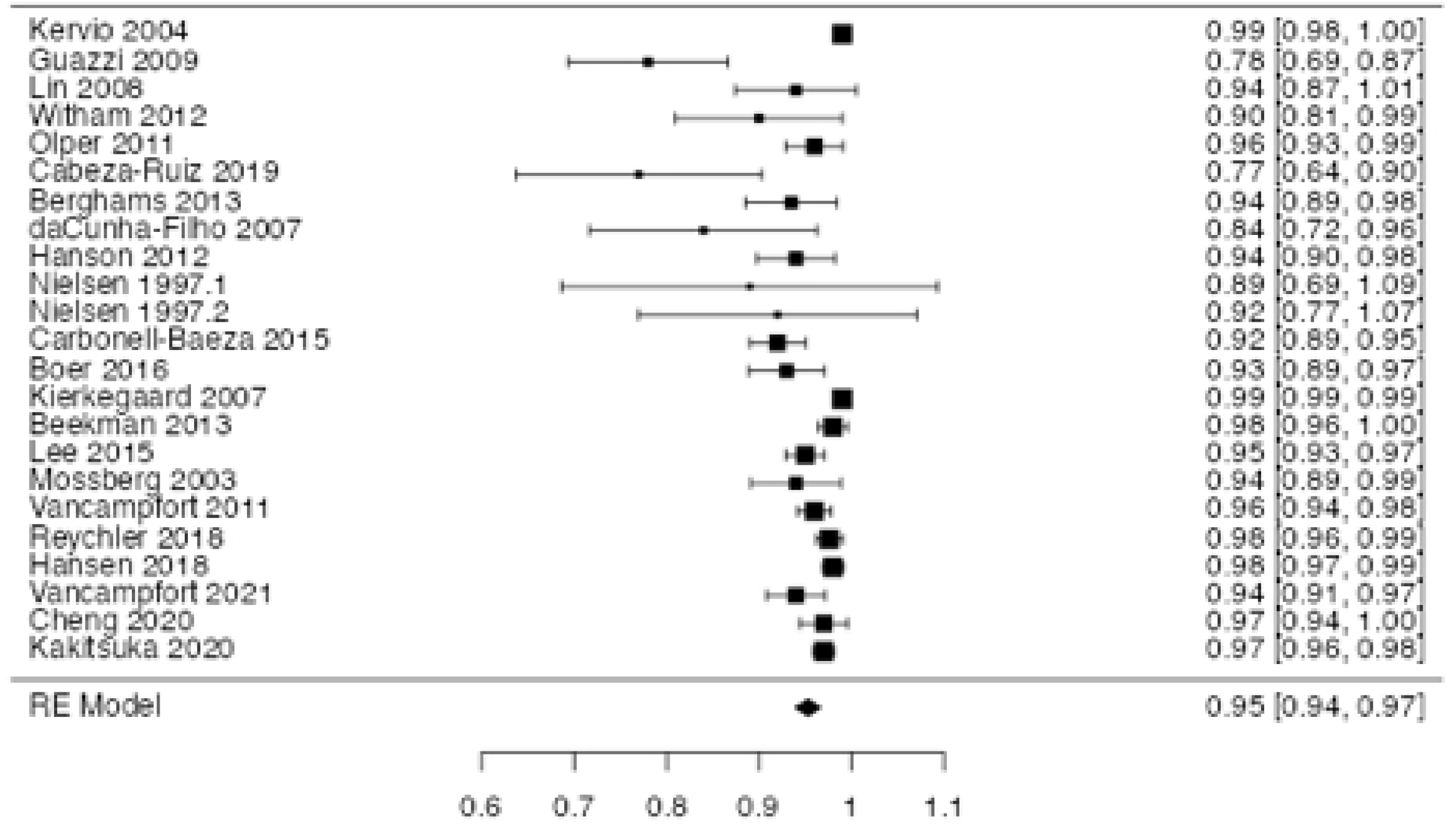
1MSTST correlation with GXT forest plot

**Fig 11:**
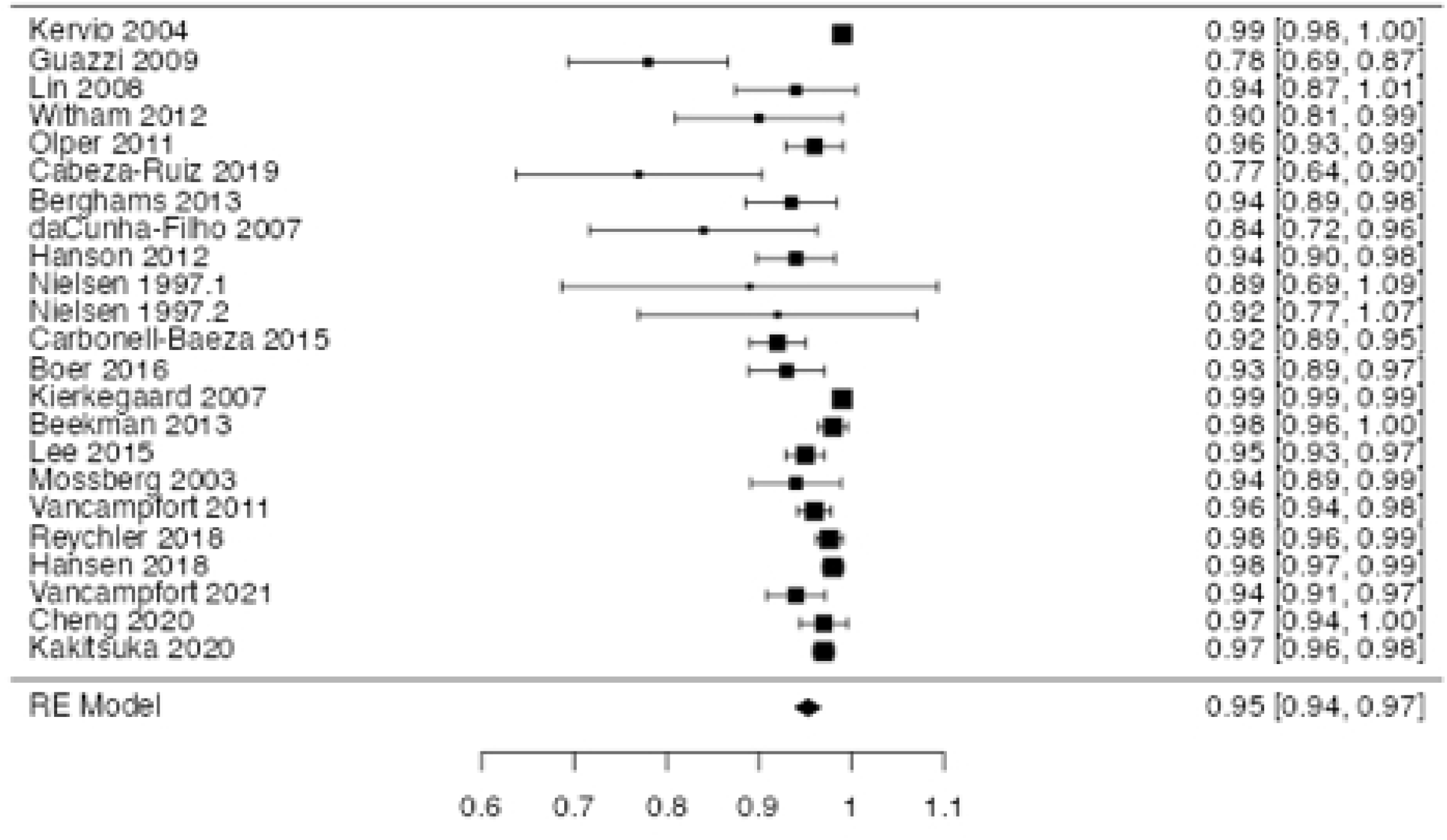
1MSTST reliability forest plot

**Fig 12:**
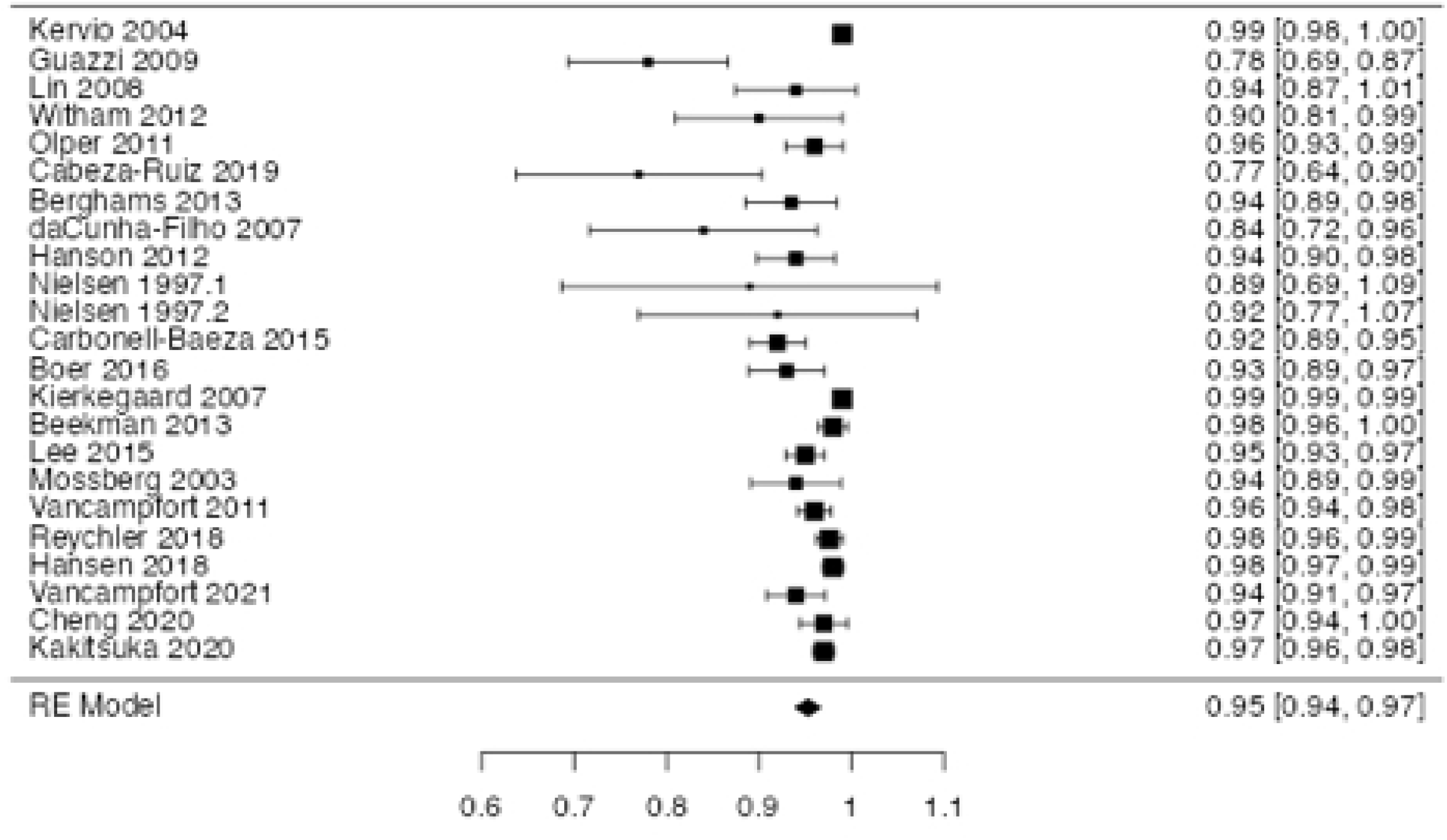
5STST reliability (ICC)

##### 3.6.2.4 2 minute step test (2MST)

One study assessed validity of the 2MST, finding a moderate correlation (r=0.54) with GXT performance, in a population of 36 Australian adults (120). The 2MST has demonstrated strong reliability (ICC>0.9) in two studies of 68 participants (121, 122)

##### 3.6.2.4 YMCA step test (YMCAST)

Kieu et al demonstrated the validity of a YMCAST equation developed and validated for a Korean population aged 19-64 years, in a population of Vietnamese participants, with a correlation of 0.8 with treadmill VO2max (123). Santo et al (124)also earlier demonstrated the validity of the YMCAST in a cohort of healthy participants via recovery heart rate correlation with treadmill GXT VO2max (124), demonstrating a correlation of 0.58 using an adjustable step height based on participant’s height which may limit clinical applicability, however remains a potentially valid predictor of CRF.

No included studies assessed the reliability of the YMCAST.

##### 3.6.2.5 Canadian aerobic fitness test (CAFT)

The CAFT was assessed in two studies describing three healthy cohorts of 60 participants in total (125, 126). Moderate correlation between CAFT performance and GXT performance was determined, r=0.645.

No test re-test reliability was established in included studies.

#### 3.6.3 Squat test**s**

##### 3.6.3.1 1 minute sit to stand test (1MSTST)

In total ten studies involving the 1MSTS were included, with three studies meeting the inclusion criteria for validity meta-analysis. Four studies of a total of 70 participants found an overall correlation of r=0.649 (98, 99, 101, 103). Radtke 2016, and Radtke 2017 studied a population with cystic fibrosis in pulmonary rehabilitation, with moderate and weak to

moderate correlation between the STS repetitions completed and VO2peak on GXT (98, 99). Gephine et al studied of a population with COPD demonstrated moderate to strong correlation between STS repetitions and VO2peak on GXT at 0.71 (101).

Five studies including 185 participants reported the reliability of the 1MSTST as strong, with an ICC on meta-analysis of 0.949 (80, 99, 100, 103, 127).

##### 3.6.3.2 30 second sit to stand test (30STST)

No study of the validity of the 30STST meeting our inclusion criteria was included.

Three studies, Hansen et al (2018) in a cohort of patients with COPD and Ozcan-Kahraman et al (2020) a cohort of patients with pulmonary hypertension, and Lázaro-Martínez et al (2022) a cohort of patients with obesity, of a total of 147 participants demonstrated strong reliability, with an ICC of 0.94 and 0.95, and r=0.91 respectively (84, 87, 128).

##### 3.6.3.3 The 5 repetition sit to stand test (5STST)

No study of the validity of the 5STST meeting our inclusion criteria was included.

Meta-analysis revealed ICC=0.91 demonstrating strong reliability. The 3 included studies demonstrated strong reliability individually. Curb et al (2006) studied 210 healthy participants, and found an ICC=0.8 (71). Jones et al (2013) studied 475 participants with COPD and found very strong reliability, an ICC of 0.97 (96). De Melo et al (2022) assessed reliability in 142 ICU patients at discharge (129).

##### 3.6.3.4 Ruffier-Dickson squat test (RDST)

One study of 40 healthy adults, performed by Guo et al found a correlation of 0.82 between a model incorporating test performance as quantified using participants’ height, sex, age and resting, immediate post-test and one minute post-test HR, and GXT VO2max (130).

Sartor et al (2016) found the RDST demonstrated good reliability in a population of 81 healthy adults (12). Reliability was calculated measuring both HRpeak (ICC = 0.86) and via the Ruffier-Dickson Index (RDI), incorporating resting, post-test and one minute post-test HR.

#### 3.6.4 Others

##### 3.6.4.1 Timed up and go test (TUGT)

No included study of the validity of the TUGT met our inclusion criteria.

Four studies including a total of 417 participants demonstrated strong reliability, with a meta-analysis ICC of 0.94 (35, 86, 87, 90). This population included health adults (Spagnuolo et al), a cohort of patients with pulmonary hypertension (Ozcan-Kahraman et al), a cohort of patients with Down syndrome (Cabeza-Ruiz et al) and a CCF cohort (Hwang et al)(35, 86, 87, 90).

##### 3.6.4.2 1RM leg press

Three studies analysing the 1RM leg press met the inclusion criteria, however none included reliability or validity statistics relevant to this review (75, 81, 131).

## 4. Discussion

The purpose of this review was to provide clinicians with information regarding the validity and reliability of submaximal tests of CRF that can be employed in a brief primary health care consultation with equipment that is readily available. This review included 143 studies of 49 clinical tests. Diverse populations of 15,670 total participants were included from studies meeting the inclusion criteria, as guided by the aims of the broad review, with a view to maintaining clinical relevance and applicability. Overall reliability of all tests included in meta-analyses were strong. Strongest validity was found for the SST, ISWT and 1MSTST on meta analysis.

### 4.1 Reliability

This review provides strong evidence for the reliability of the included clinical tests. The test-retest reliability of all included tests was high on meta-analysis. Meta-analysis revealed the 6MST was the most reliable test, however overall, all studies meta-analysed had a reliability of >0.9, and there was little difference between them. Of the studies included but in insufficient number to meta-analyse, good reliability was demonstrated for the 30STST, 5STST and RDST (12, 71, 84, 87, 96, 128, 129). The high test-retest reliability of the majority of the tests suggests they can be used to monitor changes in CRF over time as the results are evidently repeatable.

### 4.2 Validity

This study has demonstrated moderate to strong evidence regarding the efficacy of a variety of clinical tests to estimate CRF in adults. Amongst those included in meta-analysis, the Siconolfi step test demonstrated strongest validity from three studies of 138 participants of varied populations, including those with oncological and rheumatological conditions, as well as a healthy population (111, 112, 114).

Overall the walking tests, whilst the most studied, demonstrated poorer validity than the step or squat based tests. A high proportion of the included studies assess populations with generally poorer CRF as the test was intended, however the overall meta-analysis result supports the well documented ceiling effect of the 6MWT, and limiting the clinical applicability to the broader population (132, 133). This result supports that it may have more clinical utility in the rehabilitation setting, involving participants with lower VO2 peak. The ISWT demonstrated good validity and reliability on meta-analysis across five studies, and given this should be considered by clinicians (18, 134).

Step tests demonstrated good validity across studies of diverse populations, and have good clinical translation potential having to date been investigated to a lesser extent than walk tests. Of individual included studies, Sykes’ Chester step test paper demonstrates the strongest correlation with CRF(118). This may suggest that tests relying on HR measures conducted during test are superior to post/HR recovery based tests, and warrants further investigation. The three studies investigating the Siconolfi step test demonstrated highest correlation on meta-analysis, but with only 133 participants total, further investigation is warranted to further demonstrate the test’s value to the clinician(111, 112, 114). The 2MST demonstrated only moderate correlation with GXT CRF in the single included study, which may suggest there is insufficient duration to differentiate CRF levels in a well population (120).

Of the included squat tests, only one, 1MSTST, included sufficient data for meta-analysis. Seventy participants completing the 1MSTS with a moderate to strong correlation with CRF. The single papers were contrasting in their correlation with GXT, such as that of Guo et al (130)(RDST) demonstrated the RDST has good validity (0.82) using the author’s predictive model, however Diaz-Balboa’s (135) paper (30STST) demonstrated poor to moderate correlation – perhaps highlighting squat tests’ perceived limitations of relying more heavily on patients functional status, and reliance on HR recovery (130, 135).

### 4.3 Risk of bias

Overall quality of included studies was good, with high quality reporting to allow scientific replication. The vast majority of papers meta-analysed were moderate to low risk of bias, providing confidence that the results were unbiased. Many studies didn’t report or control for confounding factors, which may have influenced outcomes in some cases. In order for further adaptation of clinical tests of CRF in clinical practice, larger high-quality studies controlling confounding factors must be undertaken.

### 4.4 Strengths and limitations

The aim of this meta-analysis was to aggregate a wide array of populations and submaximal tests of CRF to maximise clinical application. Omission of studies of tests longer than 10 minutes, and requiring minimal but not the absence of equipment increases the risk of bias, and may have excluded highly relevant studies. This however allowed the goal of maintaining a clinical applicability lens to be met. The comparison between studies may have been limited by the heterogeneity of GXT protocols, as has been discussed by authors previously, however with a view to pragmatism, GXT performance was taken at face value from the included studies (136, 137). The selection criteria applied included only adult data in the analysis, and where papers included paediatric participants, adult data were extracted where possible and the study included, however in some cases the adult data could not be extracted in isolation and so that data set excluded, potentially impacting overall study results. The review included only English language papers, potentially excluding relevant studies. By design, this study included a broad population and large number of clinical tests which can be utilised in a standard primary care consultation. Further research is warranted into the applicability of valid and reliable clinical tests in specific populations.

## 5. Conclusion

The safe, cost-effective and accurate assessment and regular monitoring of CRF in the primary health care setting presents the clinician with the opportunity to personalise exercise prescription and counselling for their patient, whilst providing the patient with motivation and accountability to improve their health outcomes. This review has identified a number of submaximal tests of CRF which can be employed in a standard medical consultation. The SST and CST demonstrate their potential for clinical translation with further investigation in larger populations. The ISWT appears superior to the 6MWT and should be considered by clinicians. Based on the validity of the tests outlined, these can be used as an acceptable method of estimating VO2peak in a broad population, without the cost and access issues of formal GXT. The high test-retest reliability of the majority of the tests suggests they can be used to monitor changes in CRF over time. Further research is needed to optimise the translation of research-based exercise testing into regular clinical practice.

## Data Availability

As this is asystematicreviewand meta-analysis,all relevantdata can be foundwithinthe cited primaryarticles.Furthermore, all relevantdata for the meta-analysesare reportedwithinthe manuscript(forestplots)and its supplementaryfiles

